# Dynamic MRI versus conventional MRI for surgical planning in cervical spondylotic myelopathy: a retrospective cohort study protocol

**DOI:** 10.64898/2026.04.24.26351716

**Authors:** Shichang Yang, Yuanming Zhong, Bin Yang

## Abstract

**Introduction:** Cervical spondylotic myelopathy (CSM) surgery is frequently associated with residual neurological deficits, partly due to unrecognized dynamic spinal cord compression on conventional MRI. Current static imaging may miss position-dependent stenosis, resulting in insufficient or inappropriate decompression. This study aims to evaluate whether dynamic MRI-guided individualized surgery improves neurological outcomes compared to conventional MRI-based planning.

**Objectives:** This study aims to examine the association between dynamic MRI-guided surgical planning and neurological recovery in cervical spondylotic myelopathy, and to evaluate its role in identifying responsible segments, avoiding excessive surgery, and improving clinical outcomes.

**Methods:** This single-center retrospective cohort study will include 300 patients who underwent cervical spine surgery between January 2020 and December 2025 at the First Affiliated Hospital of Guangxi University of Chinese Medicine. Patients will be categorized into the dynamic MRI-guided group (n=150) or conventional MRI-based group (n=150) based on preoperative imaging modality. 1:1 propensity score matching will be performed using age, sex, BMI, disease duration, baseline mJOA score, and number of compressed segments. The primary outcome is the rate of improvement in the mJOA score at 6 months postoperatively. Secondary outcomes include VAS, NDI, reoperation rate, and time to first complication. Between-group comparisons will use t-tests/Mann-Whitney U tests for continuous variables, χ² tests/Fisher’s exact tests for categorical variables, and Kaplan-Meier estimates with the log-rank test for time-to-event outcomes. A two-sided P<0.05 will be considered significant. Analyses will be performed using R software (version 4.4.1). Ethical approval was obtained from the Medical Ethics Committee of the First Affiliated Hospital of Guangxi University of Chinese Medicine (Approval No. 2025-080-KY-01) from February 06, 2026 to February 05, 2027.

**Expected outcomes:** We hypothesize that dynamic MRI-guided surgical planning will improve neurological recovery and decompression accuracy in cervical spondylotic myelopathy, providing evidence for optimized preoperative imaging and precision spine surgery.

**Trial registration:** ChiCTR2600122088

## Introduction

CSM is caused by cervical spinal stenosis and age-related cervical degenerative changes, including degenerative intervertebral disc herniation, osteophyte formation, ligamentum flavum hypertrophy, and ossification of the posterior longitudinal ligament, which lead to persistent cervical spinal cord compression and subsequent spinal cord conduction dysfunction [1]. Cervical segmental instability, minor trauma, or repetitive strain can further aggravate spinal cord injury and accelerate disease progression [2].

As the most common non-traumatic cause of spinal cord dysfunction in middle-aged and elderly populations, CSM has become a major public health concern in global spinal surgery. In recent years, driven by population aging, lifestyle changes, and advances in diagnosis and treatment, global prevalence and surgical rates for CSM have risen consistently [3].

In terms of geographical distribution, this disease shows distinct racial and regional variations. East Asian populations have a higher prevalence of developmental cervical spinal stenosis and more frequent ossification of the posterior longitudinal ligament, leading to significantly higher overall incidence and clinical presentation rates of CSM than in European and American Caucasian populations [4].

At present, surgical intervention is the main treatment for CSM, and most patients achieve certain neurological function improvement after surgery [3, 5]. However, some patients still have inadequate spinal cord decompression postoperatively, characterized by persistent neck pain, poor neurological function recovery, and even symptom recurrence [6].

Furthermore, dynamic spinal cord compression is often overlooked because the cervical spine is highly mobile, and most MRI examinations are routinely performed in a single neutral position [7]. Spinal cord compression in some CSM patients is position-dependent: mild or absent on static neutral-position MRI, but distinct in cervical hyperextension or flexion [7, 8]. This may cause inaccurate preoperative localization of responsible segments and inadequate surgical decompression, impairing postoperative neurological recovery and even leading to persistent or recurrent symptoms [9].

Cervical dynamic MRI identifies occult spinal stenosis not visualized on static imaging by assessing spinal cord morphology and compressive changes during active hyperextension and hyperflexion, enabling comprehensive evaluation of compressed segment severity and distribution. It also uncovers pathological changes undetectable on neutral-position MRI, thereby informing CSM surgical strategy formulation [10, 11].

Data for this study will be derived from routine clinical practice between 2020 and 2025. To minimize concerns regarding data dredging and selective reporting, we developed and published a detailed analytical plan prior to accessing the full dataset for hypothesis testing, and registered the study protocol to improve transparency and reproducibility of retrospective results. Accordingly, this study aims to evaluate dynamic spinal cord compression using dynamic cervical MRI and establish individualized surgical strategies, enabling precise localization of responsible segments and targeted decompression. This approach overcomes the diagnostic limitations of conventional static MRI, reduces overtreatment and iatrogenic injury, enhances the accuracy and safety of surgery for cervical spondylotic myelopathy (CSM), and improves neurological outcomes and quality of life in patients. As a promising precision management strategy in the spine surgical care of CSM, it carries important clinical value and practical necessity.

## Methods and analysis

### Study Design and Setting

This study protocol is designed and reported in accordance with the Strengthening the Reporting of Observational Studies in Epidemiology (STROBE) Statement for cohort studies and the STROBE-Protocol guidelines.

This is a single-center, retrospective cohort study that will use routinely collected clinical data. The study will be conducted at the Department of Spinal Orthopaedics, the First Affiliated Hospital of Guangxi University of Chinese Medicine. The clinical and imaging data of patients who underwent cervical spine surgery in our hospital from January 1, 2020, to December 31, 2025, were retrospectively extracted from the hospital’s electronic medical record system and imaging files.

A retrospective cohort design is adopted for the following reasons: dynamic cervical MRI has been gradually implemented in routine clinical practice, forming natural exposure groups. This design allows large-sample and long-term follow-up data to be obtained efficiently for evaluating surgical efficacy and safety. To minimize selection bias, strict inclusion and exclusion criteria will be applied, and 1:1 propensity score matching (PSM) will be performed to balance confounding factors between groups.

### Study Status

No prospective patient recruitment will be performed. Data have not yet been extracted for the study and will not be collected until the protocol is approved. The index date for all patients was defined as the date of surgery. Subsequent data collection is expected to be completed by August 2026.The planned schedule of enrolment, interventions, and assessments is presented in Figure 1, according to the SPIRIT 2025 statement.

**Figure 1.**
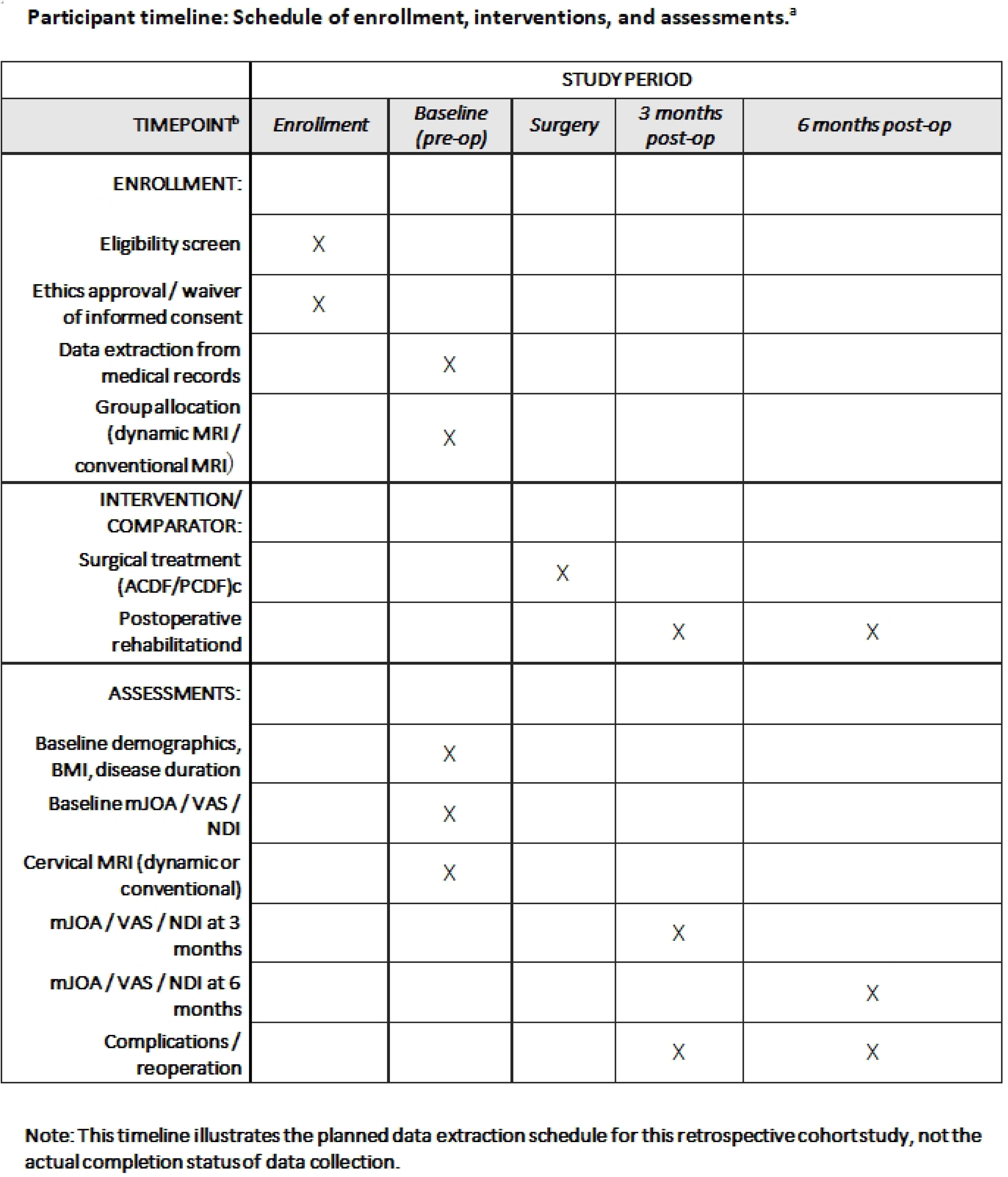
SPIRIT 2025 schedule of enrolment, interventions, and assessments.

### Objectives

#### Primary Objective

To evaluate whether preoperative dynamic MRI-guided individualized surgical planning improves the 6-month postoperative modified Japanese Orthopaedic Association (mJOA) score improvement rate compared with conventional static MRI-based planning in patients with cervical spondylotic myelopathy (CSM).

#### Secondary Objectives

1. To compare postoperative improvements in Visual Analogue Scale (VAS) and Neck Disability Index (NDI) scores at 3 and 6 months between the dynamic MRI-guided group and the conventional MRI-based group.
2. To explore the value of dynamic MRI in detecting occult dynamic spinal cord compression and its impact on modifying surgical strategies (e.g., changing the number of decompression levels or surgical approach selection).
3. To identify demographic, clinical, and imaging factors associated with neurological recovery after CSM surgery, including age, disease duration, baseline severity (mJOA score), number of compressed segments, and presence of dynamic compression.

### Study Participants

This study will conduct a retrospective analysis of 300 patients diagnosed with cervical spondylotic myelopathy (CSM) who received surgical intervention at the Department of Spinal Orthopedics and Traumatology, the First Affiliated Hospital of Guangxi University of Chinese Medicine, between January 2020 and December 2025. Eligible cases will be identified from the existing clinical database in accordance with the established inclusion criteria. Details of patient screening, enrollment, grouping, and follow-up are presented in Fig. 2.

**Figure 2.**
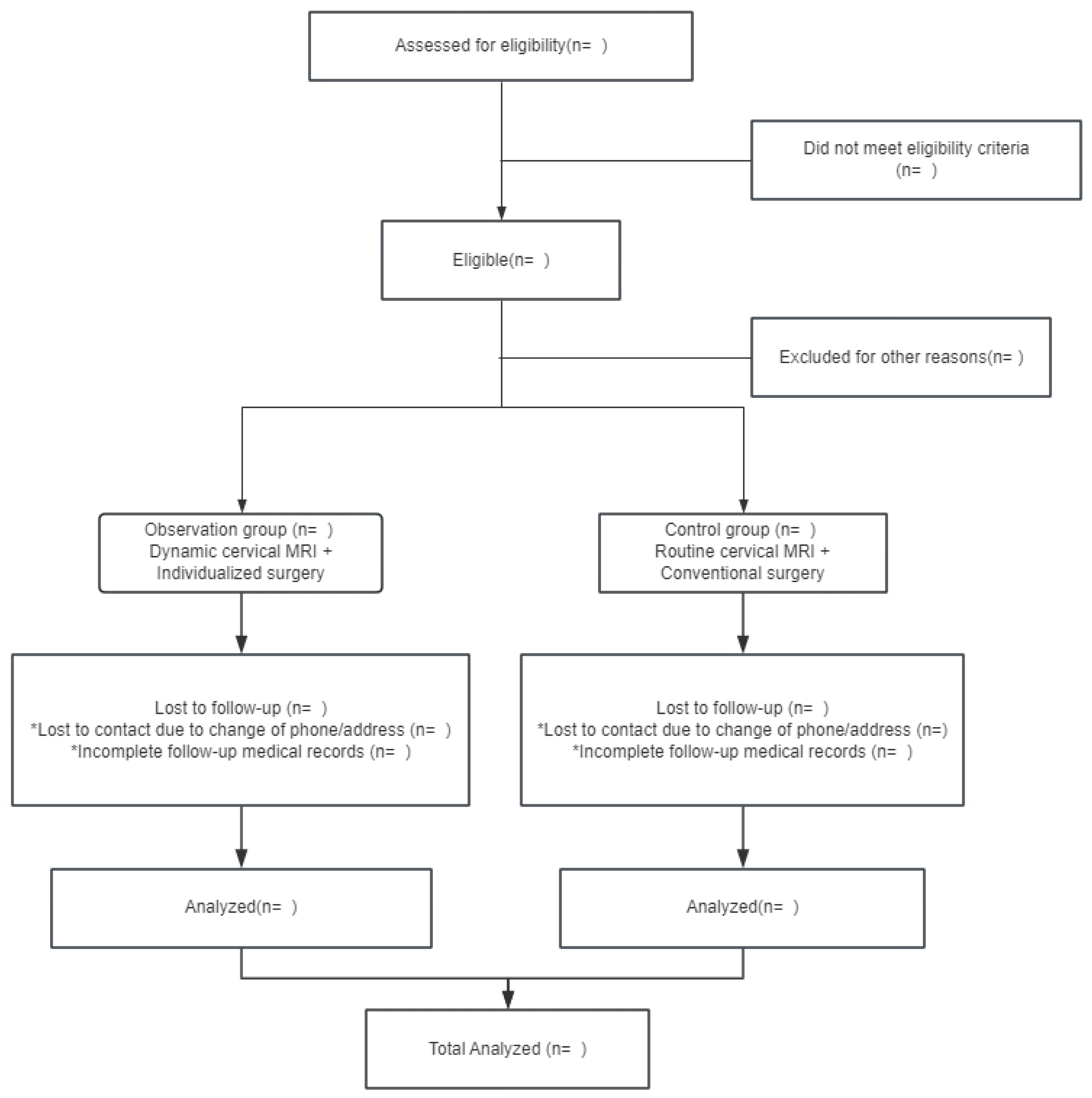
Flowchart of patient screening, enrollment, grouping, and follow-up.

### Inclusion Criteria

1. Aged 18–60 years, regardless of gender.
2. Disease duration > 3 months and ineffective conservative treatment.
3. Availability of preoperative MRI (dynamic or conventional) and minimum 6-month postoperative follow-up data.

### Exclusion Criteria

1. Pregnant or lactating women.
2. History of severe cervical spine trauma, cervical spine surgery, or other spinal surgery.
3. Preoperative use of analgesic medications.

### Grouping strategy

Patients will be assigned to two groups based on the preoperative imaging modality actually utilized in routine clinical practice, as documented in medical records: an observation group (undergoing preoperative dynamic cervical MRI) and a control group (undergoing preoperative conventional neutral position cervical MRI). To balance baseline characteristics, including age, sex, BMI, disease duration, baseline mJOA score, and number of compressed spinal segments, 1:1 propensity score matching (PSM) will be performed. Outcome assessors will be blinded to group allocation during follow-up.

Patients in the observation group underwent preoperative dynamic cervical MRI scans using a 3.0 T scanner. Sequences will include T2-weighted sagittal images obtained in the neutral position, 30° flexion, and 20° extension. An individualized surgical plan was formulated jointly by two radiologists and two spinal surgeons at the associate professor level or above, based on the number of compressed segments, site of compression, cervical spine stability, and other related factors. Anterior cervical discectomy and fusion (ACDF) will be performed for patients with anterior compression of 1–2 segments. Posterior cervical decompression and fusion (PCDF) will be adopted for patients with compression of ≥3 segments or posterior spinal cord compression. All surgeries were performed by chief physicians or associate professors with at least 10 years of clinical experience, and intraoperative neurophysiological monitoring will be routinely applied. For ACDF, complete anterior decompression of the spinal cord and nerve roots will be achieved by discectomy, excision of the herniated nucleus pulposus, resection of the posterior longitudinal ligament, and removal of osteophytes. Interbody fusion will be conducted using appropriately sized cervical cages filled with autologous bone, artificial bone, or bone graft substitutes, with rigid internal fixation to maintain cervical alignment. For PCDF, spinal cord decompression will be performed via posterior laminectomy or laminoplasty, followed by posterior pedicle screw fixation and fusion. Standardized stepwise rehabilitation was initiated on postoperative day 3 under the guidance of specialized rehabilitation physicians.

Patients in the control group will only undergo conventional preoperative neutral? position cervical MRI, based on which the surgical plan will be determined. The criteria for surgical approach selection, decompression techniques, internal fixation and fusion strategies, and postoperative rehabilitation protocol will be identical to those applied in the observation group.

### Outcome Measures

The primary outcome will be the modified Japanese Orthopaedic Association (mJOA) score, which will serve as the core evaluation index. The clinical efficacy of individualized surgical diagnosis and treatment guided by dynamic cervical MRI for neurological function recovery will be evaluated using mJOA scores at 3 and 6 months postoperatively, as well as the improvement rate in mJOA scores. An mJOA improvement rate ≥75% will be defined as excellent, 50%–74% as good, 25%–49% as fair, and <25% as poor.

The secondary outcomes included the Visual Analogue Scale (VAS) and Neck Disability Index (NDI), assessed at 3 and 6 months postoperatively.

### Data Collection and Management

As this is a retrospective study, no prospective patient contact or intervention will be performed. All data will be extracted from existing medical records and imaging archives. All collected data will be de-identified to protect participants’ privacy; personal identifiers (such as name, ID number, telephone number, and medical record number) will be removed or encrypted, and only anonymous research codes will be used for data management and analysis throughout the study.

Two trained researchers will independently extract and cross-check the data before entering it into a dedicated database. Baseline data (demographics, clinical data, conventional cervical MRI, preoperative mJOA/VAS/NDI scores) will be extracted from preoperative records; postoperative follow-up data (3 and 6 months) will be collected from outpatient and imaging records, with missing data supplemented by telephone follow-up and electronic health record review.

All data will be recorded in standardized CRFs, then double-independently entered into Excel and cross-validated. An internal quality control process, supervised by the ethics committee and led by the principal investigator, will be implemented for regular random checks to ensure data integrity and protocol compliance.

## Statistical Analysis

### Data processing and analysis

After data cleaning, the application of overall and individual exclusion criteria (see Inclusion and Exclusion Criteria) will be documented, and the final size of the analytic study population will be determined. Normality of data distribution will be assessed visually and by calculating skewness and kurtosis. Normally distributed data will be presented as mean ± standard deviation (SD), whereas non-normally distributed data will be reported as median with interquartile range [M (IQR)].

Descriptive statistics will summarize the demographic and clinical characteristics of the study population, presented for the entire cohort and stratified by imaging modality (dynamic MRI group: preoperative neutral plus extension–flexion MRI; conventional MRI group: preoperative neutral MRI only) and severity of myelopathy (mJOA ≥ 15: mild; mJOA 12–14: moderate; mJOA < 12: severe). In between-group comparisons of baseline characteristics, categorical variables will be analyzed using the Pearson χ² test or Fisher’s exact test, and continuous variables will be compared using the independent samples t-test or Mann–Whitney U test according to data normality. All statistical analyses will be performed using R software (version 4.4.1).

### Primary Outcome Analysis

The primary outcome will be the improvement rate of the mJOA score at 6 months after surgery, calculated using the formula: [(postoperative mJOA score - preoperative mJOA score) / (17 - preoperative mJOA score)] × 100%. Patients will be stratified into mild, moderate, and severe groups according to their baseline mJOA scores. Preoperative outcomes among the three groups will be compared using one-way analysis of variance (ANOVA) or the Kruskal-Wallis test. A Bonferroni post-hoc test will be performed if P < 0.1, and the moderate and severe groups will be merged if necessary. Baseline characteristics (including gender, age, and BMI), mJOA score improvement rates, and efficacy grades will be compared between the two groups using the corresponding statistical tests. For binary efficacy outcomes, the Pearson χ² test will be used to calculate the relative risk (RR) and 95% confidence interval (95% CI).

### Secondary Outcome Analysis

Secondary outcomes (Neck Disability Index [NDI] and Visual Analog Scale [VAS] scores) will be analyzed using a linear mixed-effects model to evaluate changes over time, with adjustments for age and baseline mJOA score, and a random intercept term. Normality of residuals will be assessed using Q-Q plots. Pairwise comparisons will be performed with Bonferroni correction, and the results will be presented as estimated values with 95% CIs.

### Subgroup Analysis of the Dynamic MRI Group

In the subgroup analysis of the dynamic MRI group, the rate of surgical strategy change and its 95% CI will be calculated, and baseline differences between subgroups will be compared. The composition ratio will be calculated according to the body position where the compressive lesion will be detected. Cochran’s Q test will be used to compare the detection rate of compressive lesions among different body positions, and then the mJOA score improvement rate will be compared between the group with newly detected compression in dynamic positions and the group with only static compression.

### Multivariate Analysis

Multiple linear regression will be used to evaluate the independent effect of dynamic MRI on the mJOA score improvement rate (after adjusting for relevant covariates). Binary logistic regression will be performed to analyze the association between the detection of occult compression by dynamic MRI and an "excellent" postoperative outcome, and the odds ratio (OR) and 95% CI will be calculated.

### Sample size calculation

A two-sided test will be adopted based on previous studies on surgical outcomes for cervical spondylotic myelopathy. The type I error (α) will be set at 0.05 with a statistical power of 0.80. The sample size estimate will be based on expected between-group differences in primary outcomes: the improvement rate of the modified Japanese Orthopaedic Association (mJOA) score and postoperative complication incidence. The minimum required sample size will be 120 patients per group. To account for a 20% loss to follow-up or exclusion, 150 patients will be enrolled in each group. The total sample size will be 300 cases.

### Missing Data Handling

Analyses will follow the intention-to-treat principle. For primary outcomes, missing mJOA scores will be handled by multiple imputation; complete-case analysis will be used only if the missing rate <5% and missingness is random. Missing adverse events will be verified via medical records; unverifiable data will be excluded with reasons documented.

### Research Schedule

Data extraction from historical medical records (2020-2025) has not yet commenced and is scheduled to begin upon protocol acceptance; it is expected to be completed by August 2026. No statistical analyses for the predefined outcomes have been performed to date. The entire study, including completion of follow-up data collection, statistical analysis, and manuscript preparation, is scheduled to be finalized by the end of January 2027.

### Patient and Public Involvement

No patients or public will be involved in the design of this retrospective study.

### Ethics Approval

This study was approved by the Medical Ethics Committee of the First Affiliated Hospital of Guangxi University of Chinese Medicine between January 2026 and January 2027 (Ethics Approval No. 2025-080-KY-01). Since this is a retrospective cohort study involving only the review of existing medical records without direct contact with participants or interference with their clinical management, the requirement for informed consent was waived by the Institutional Review Board (IRB)/Ethics Committee.

### Data Availability

No datasets have been generated or analyzed at the time of protocol submission. Data extraction for this study has not yet commenced. Data will be made available from the corresponding author upon reasonable request after study completion, subject to ethical approval.

## Discussion

This study is a single-center retrospective cohort study designed to evaluate the value of individualized surgical planning for cervical spondylotic myelopathy (CSM) under the guidance of cervical dynamic MRI. To date, few studies have investigated the use of cervical dynamic MRI in the preoperative assessment of the responsible compressive segments in CSM. Clinical practice relies primarily on conventional neutral-position MRI, which often fails to detect dynamic spinal cord compression associated with positional changes. This may lead to misidentification of the responsible segments, resulting in insufficient decompression or excessive surgery [7, 12, 13]. To examine whether dynamic MRI can address this clinical challenge, the present study directly compared the clinical outcomes of surgical planning based on dynamic MRI versus conventional MRI to determine whether it improves surgical accuracy, enhances neurological recovery, and reduces the risk of unnecessary surgery.

This study will have a rigorous design, with selection and information bias minimized via strict inclusion/exclusion criteria and double data verification. Sample size will be calculated based on the modified JOA score (α=0.05, 1-β=0.80), with 100 patients in each group to ensure statistical power. Efficacy will be evaluated multidimensionally using modified JOA, VAS, and NDI scores. The observation group underwent dynamic MRI (neutral, 30° flexion, 20° extension) to accurately assess dynamic spinal cord compression. Perioperative, surgical, and rehabilitation protocols will be identical between groups, with only imaging evaluation differing, ensuring clear result attribution.

The use of dynamic MRI was based on the pathological mechanism of position-related spinal cord compression in CSM, which is closely associated with cervical degenerative segmental instability. Hyperextension exacerbates compression via ligamentum flavum folding, osteophytes, and posterior longitudinal ligament ossification, while hyperflexion worsens it via disc herniation, vertebral displacement, and reduced spinal canal sagittal diameter [14, 15]. Clinically, patients often have position-dependent neurological symptoms, exacerbated by cervical hyperextension/flexion and relieved in the neutral position [16]. Dynamic MRI, particularly in hyperextension, outperforms other imaging modalities in capturing this dynamic process, identifying conventional MRI-undetectable potential compression to distinguish static degeneration from activity-aggravated compression [17, 18]. This imaging evidence enables individualized surgical plans to achieve accurate, responsible-segment decompression, avoid ineffective surgery, reduce iatrogenic injury and postoperative complications, and ultimately improve neurological function, pain, and cervical function [19, 20].

Limitations of this study are as follows. First, the retrospective, single-center design inherently has selection bias, and geographical/ethnic differences in the epidemiology of CSM limit the generalizability of our findings due to center-specific patient populations and surgical practices. Second, dynamic MRI increases patients’ cost and time burden, and its cost-effectiveness restricts its widespread application in resource-limited settings. Third, dynamic MRI poses safety risks for patients with severe CSM: cervical hyperextension may exacerbate spinal cord compression and neurological deterioration, making severe cases (obvious spinal stenosis, segmental instability, or T2 hyperintensity on static MRI) unable to undergo safe examination and introducing selection bias; close monitoring is required, with immediate termination of the examination if neurological symptoms worsen. Fourth, non-standardized cervical movement (due to individual differences in pain tolerance/neurological deficits) and supine positioning (failing to replicate the physiological spinal load in the upright position) may reduce imaging consistency.

If this study confirms that the dynamic MRI-guided individualized surgical protocol has significant clinical advantages, it will support the inclusion of this protocol in the standardized preoperative evaluation system for CSM. This will promote the transformation of CSM diagnosis and treatment from conventional empirical surgery to precision surgery, improve the efficiency of neurological function recovery, optimize the surgical planning process, and ultimately reduce the medical and rehabilitation burden on patients.

### Protocol Amendments

Any amendments to this protocol will be documented with the date, description of changes, and rationale. Substantial modifications affecting study design, outcomes, or patient safety will be submitted to the Medical Ethics Committee of the First Affiliated Hospital of Guangxi University of Chinese Medicine for approval prior to implementation, and updated on the Chinese Clinical Trial Registry (ChiCTR2600122088).

## Data Availability

For Study Protocols: No datasets were generated or analysed during the current study. All relevant data from this study will be made available upon study completion.

